# Environmental detection of parasites in the marginalized Paiute reservations compared to a nearby area

**DOI:** 10.1101/2023.10.24.23297407

**Authors:** Shannon McKim, Kristen Kopystynsky, Nathaniel Wolf, Fahim A. Akbar, Maria Elena Bottazzi, Peter J. Hotez, Rojelio Mejia

## Abstract

The amounts of parasite DNA in soil samples from different playgrounds and other public areas can help identify areas of possible microbe transmission as well as giving indications of possible occurrence of parasite infection in nearby communities. We collected 207 soil samples from parks located on Paiute indigenous tribal areas in southwest Utah and from higher income city of St. George, Utah, and tested them for the presence of 11 parasites that can cause human disease. Molecular tests revealed elevated odds ratio of detecting the helminth *Trichuris trichiura* 3.072 (1.114 to 8.065) and any protozoa (not including *Acanthamoeba*) 3.036 (1.101 to 7.966) in the tribal land playgrounds compared to St. George parks. These findings support previous studies showing that areas in lower socioeconomic communities, especially marginalized communities, tend to have higher presence of parasites in the soil that may lead to higher rates of disease prevalence.

## Text

Parasites and parasite larvae in soil pose a potentially serious health risk to those living in marginalized communities. The transmission of parasites to humans can lead to several serious health problems ranging from mild allergies to fatal amoebic encephalitis. Children are particularly vulnerable to fecal oral transmission from soil contaminated by zoonotic and human derived parasites. Protozoans, (e.g *Giardia intestinalis*) and helminths (e.g *Trichuris trichiura*) have complex life cycles and can remain dormant for extended periods in soil and water, exposing humans to these transmission risks.

While parasitic transmission can be blunted through improved sanitation efforts, clean drinking water, and access to medical care, many underserved populations struggle to meet these necessities, ^1–3^ and as a result, have been established to have a higher seroprevalence of parasitic infections^4^. Children in these populations are of particular concern because of the increased hand-to-mouth transmission from soil^5^. Additionally, early health disadvantages on a child can have a corrosive effect on an individual’s success later in life, long after the original illness has resolved^6^.

It is well-established that indigenous reservations generally experience greater rates of poverty and socioeconomic disadvantages^7^. A link between a higher burden of soil parasites in lower SES populations has been established in other areas of the country, so it is reasonable to suspect a similar trend in Native Lands^8^. To our knowledge, soil in the native Paiute lands in southern Utah has not been tested for the presence of parasites. In this study, we analyzed soil samples collected from various parks and public spaces located within the Paiute tribal band lands. We then compared the parasite presence in these samples with those collected from St. George, Utah areas with similar communal usage. Our goal, with this research, is to raise awareness regarding parasitic transmission, and analyze differences in the number of organisms found between regions of differing socioeconomic status.

Written permission to collect soil samples was obtained from The Paiute Indian Tribe of Utah Tribal Council and the Parks Division of the city of St. George. Soil samples were collected from publicly used spaces in the St. George area and in Paiute native lands. The Utah and Arizona locations collected from are generally desert habitats or other similarly dry environments. The samples were collected during a 12 month period in 2021. Sampling locations in St. George parks were defined by higher socioeconomic status. A total of 207 soil samples were collected via 50 mL test tubes and stored in airtight plastic storage containers. Soil samples collected were then stored at 4° C on Rocky Vista University campus. Samples were photographed geo-tagged with the exact location from where they were collected. Soil samples were moved and processed at Baylor College of Medicine where they were stored at –20 C prior to further investigative methods. Soil samples were weighed, and dry weight was recorded. Samples were then transferred to a new Falcon tube with no more than 25mL of solid material per tube. If there were more than 25 mL of soil available, then the sample was divided evenly between two separate Falcon tubes. 0.05% TWEEN/PBS was added to each sample until total volume of each tube measured 45 mL. Samples were then centrifuged at 1500 RPM for 5 minutes. Supernatants were then filtered using a 3.0 um SSWP membrane (Milipore, Tullagreen, Ireland). Membranes were processed using FastDNA SPIN Kit for Soil (MP Biomedicals, Santa Clara, California, USA) and were heated at 90° C for 10 minutes prior to bead beating. DNA was then analyzed further through a multi-parallel quantitative polymerase chain reaction (qPCR) to screen for 11 different parasitic-specific DNA sequences (*Ancylostoma duodenale, Ascaris lumbricoides, Necator americanus, Strongyloides stercoralis, Toxocara canis/cati, Trichuris trichiura, Acanthamoeba* species, *Blastocystis species, Cryptosporidium species, Entamoeba histolytica, Giardia intestinalis*) using primer and probes from a previous study^9^. *Acanthamoeba* control DNA came from *Acanthamoeba castellanii* (ATCC strain 30010) ^10^. Primer and probes for *Acanthamoeba* species include:

Forward Primer: CCCAGATCGTTTACCGTGAA

Reverse Primer: TAAATATTAATGCCCCCAACTATCC

Probe 5’FAM MGB: CTGCCACCGAATACATTAGCATGG^11^

The percentage of population living in poverty (as defined by the U.S. Census Bureau, 2020) on Paiute Reservations in Utah was 28.1% ^12^, compared to 10.5% in St. George^13^, a 2.6 times higher increase. For children under 18 years old, the poverty rate in the Paiute Reservation was 49.4% compared to 8% in St. George, an increase of 6.2 times higher poverty rate (Figure 1).

**Figure 1.**
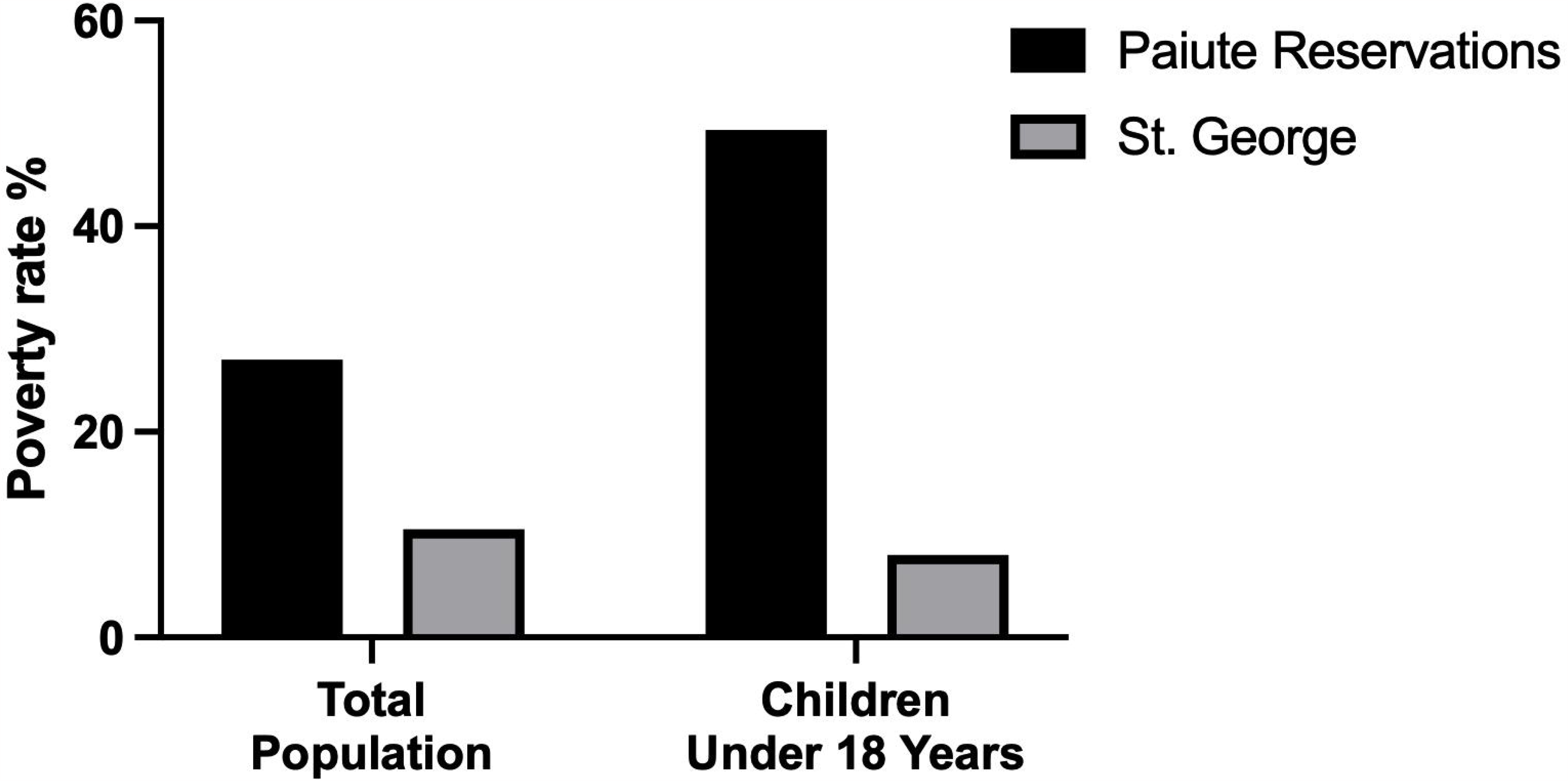
The poverty rate in Paiute Reservations compared to St. George.

Parasites detected in the Paiute reservation soils included 15 samples with helminths compared to 7 samples from St. George (Table 1).

**Table 1.**
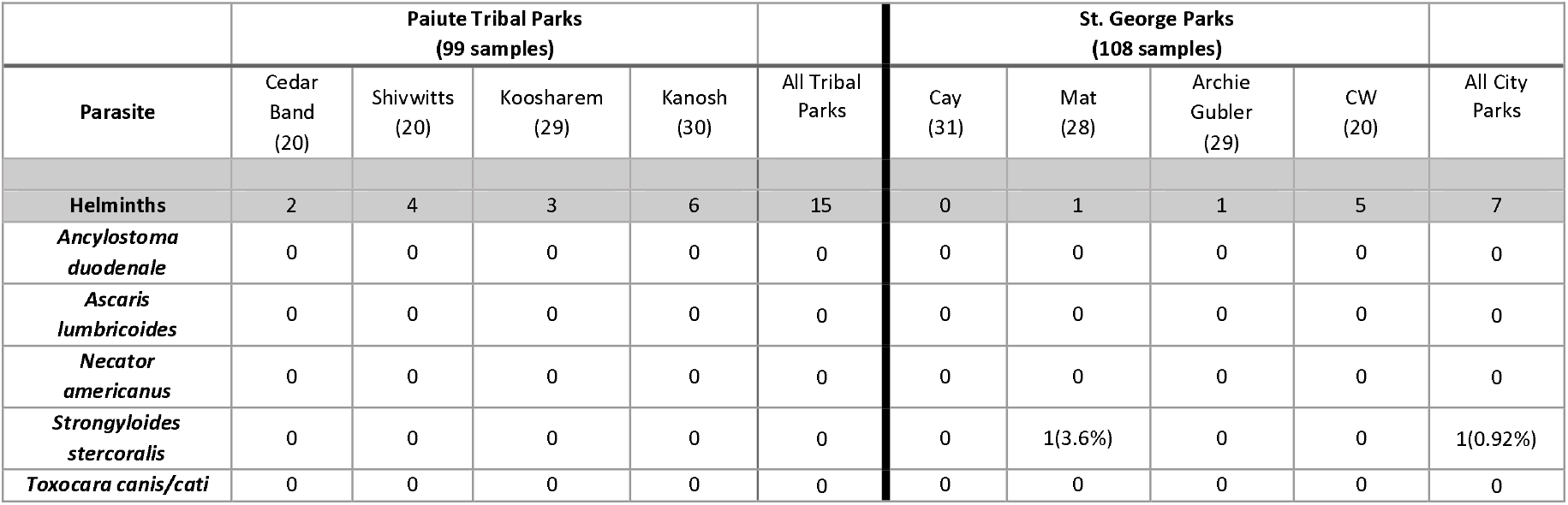
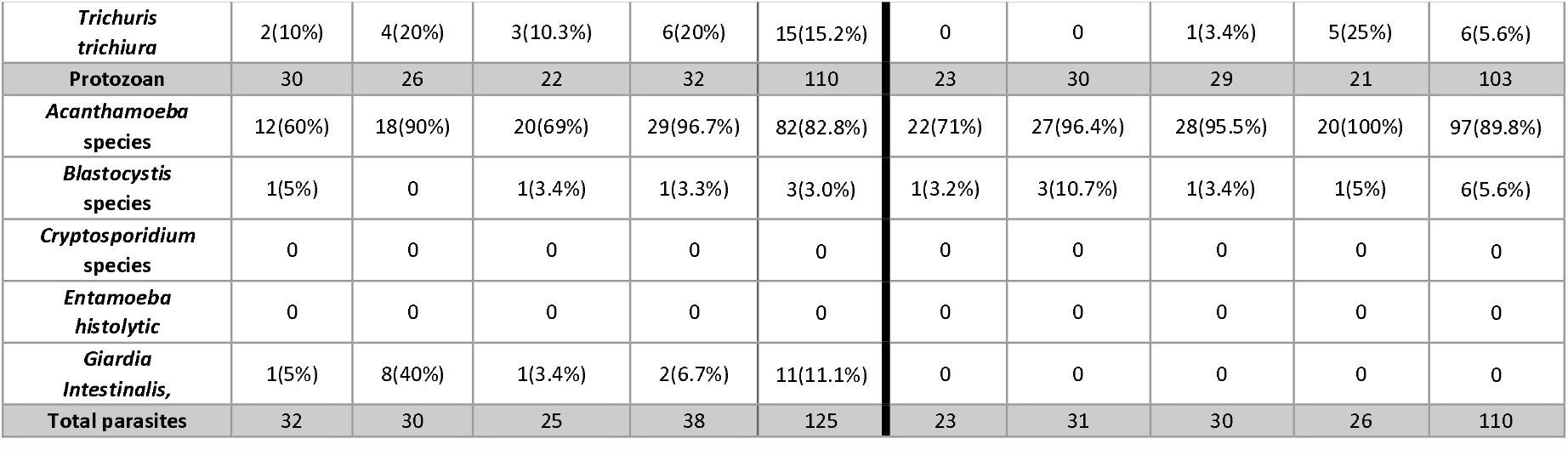
Prevalence rates of parasites found in soil samples. Soil types collected a. dirt, b. rocks, c. sand, d. woodchips.

**Table 2.** Odds-Ratio Analysis.

| **Parasite** | **Paiute Tribal Parks** | **St. George Parks** | **Odds Ratio** | **P-value** |
| --- | --- | --- | --- | --- |
| **Helminths** | 15 | 7 | 2.608<br>(1.006 to 6.637) | *0.0406 |
| *Strongyloides stercoralis* | 0 | 1 | N/A | N/A |
| *Trichuris trichiura* | 15 | 6 | 3.072<br>(1.114 to 8.065) | *0.0209 |
| **All Protozoan** | 94 | 103 | 0.9126<br>(0.2872 to 2.902) | 0.8878 |
| **Protozoan not *Acanthamoeba*** | 15 | 6 | 3.036<br>(1.101 to 7.966) | *0.0224 |
| *Acanthamoeba* species | 79 | 97 | 0.4479<br>(0.2076 to 0.9744) | *0.0436 |
| *Blastocystis* species | 3 | 6 | 0.5313<br>(0.1428 to 2.2029) | 0.3735 |
| *Giardia Intestinalis*, | 12 | 0 | N/A | N/A |
| **Total parasites** | 109 | 110 | N/A | N/A |
| **Total parasites not *Acanthamoeba*** | 30 | 13 | 3.177<br>(1.589 to 6.256) | *0.0012 |

In the Paiute tribal parks there was a 3.072 (1.114 to 8.065) odds ratio for detecting *Trichuris trichiura* compared to St. George parks (Table 3).

Our study found a considerable number of parasites in public parks in the southwest Utah region, and for the first-time characterized parasites present on Paiute Native lands. Furthermore, our data demonstrates a high prevalence of parasites that are relevant to public health concerns, including *Trichuris trichiura* and *Giardia intestinalis*. While *Acanthamoeba* species was found in more samples from the St. George parks there was no difference in concentration of DNA. However, *Acanthamoeba* is known to be a free-living organism found in a very vast array of environments. Although it is an important consideration as an infectious agent itself or as a vector for other infectious agents, its presence does not necessarily reflect the effects of poverty. When this ubiquitous organism was not included in our analysis, the higher all-parasite burden of Paiute parks becomes rather clear (OR 3.177, P-value 0.0012).

A previous study demonstrated a significant association between *Toxocara catis* and lower socioeconomic status in various NYC boroughs^8^. Our findings are similar in demonstrating a higher parasite burden in the lower socioeconomic areas, which may be adding to the already notable health disparities between native reservations and the rest of the United States.

One major difference which may contribute to the higher rates of parasites in tribal areas is lack of fencing in the tribal parks, as opposed to the St. George parks which were fenced; increased fencing could be an important measure that would decrease soil-borne parasites. Tribal elders as well as St. George city officials should be encouraged to initiate interventions including measures to increase awareness and surveillance among local healthcare workers of *Trichuris* and *Giardia* (which often go undiagnosed) as well as *Acanthamoeba*, which although it rarely causes infection can facilitate other pathogens; measures to control population of stray dogs and feral cats; and increased sanitation measures within the parks. Educational programs should include handwashing and encouraging children not to put foreign objects in their mouth or touch the soil with their face.

As a caveat, our study was limited by the precise nature of the qPCR probes used, which are specific to human parasites. However, many zoonotic parasites have not yet been sequenced to the degree of detail needed to differentiate them from other species, and therefore could have been cross detected. For example, *Trichuris vulpis*, a species of whipworm known to infect dogs, has been known to cause false detections with *T. trichiura* qPCR probes^14^. Additionally, our study was limited to the species we happened to test for with our qPCR probes. In addition, the possibility of human error must be considered along with randomization methods of soil retrieval. Soil samples were selected at random however specific soil densities and moisture levels were not standardized; furthermore, future studies could include animal surveillance to increase understanding of study results as well as the consideration of factors such as the extent of vegetation and the frequency of non-precipitation watering in each park.

While important, soil studies are limited in their ability to detect actual transmission and infection rates in a population. Further research will be necessary to establish seroprevalence of parasites in Paiute populations vs. other local populations. This is necessary to establish whether this increased parasite burden leads to increased infection rates.

## Data Availability

All data produced in the present study are available upon reasonable request to the authors

## Acknowledgments

We wish to thank the Paiute Indian Tribe of Utah Tribal Council for their support and approval of this project. Also special thanks to Jackson Been and Matthew Gaskins for helping to collect samples.

## References

1. Lakhani N., 2021. Tribes without clean water demand an end to decades of US government neglect. 2021. Available at: https://www.theguardian.com/us-news/2021/apr/28/indigenous-americans-drinking-water-navajo-nation. Accessed August 31, 2023.

2. Hubbard S, Chen PM., 2022. Falling Short: Examining Medical Debt and Cost Avoidance in American Indian and Alaska Native Households. J Health Care Poor Underserved 33

3. Indian Health Service., 2023. Safe Water and Waste Disposal Facilities Fact Sheets. Available at: https://www.ihs.gov/newsroom/factsheets/safewater/. Accessed. 2023

4. Won KY, Kruszon-Moran D, Schantz PM, Jones JL., 2008. National seroprevalence and risk factors for zoonotic Toxocara spp. infection. American Journal of Tropical Medicine and Hygiene 79

5. Kassaw MW, Abebe AM, Tlaye KG, Zemariam AB, Abate BB., 2019. Prevalence and risk factors of intestinal parasitic infestations among preschool children in Sekota town, Waghimra zone, Ethiopia. BMC Pediatr 19: 1–10

6. Braveman P, Barclay C., 2009. Health disparities beginning in childhood: a life-course perspective. Pediatrics 124 Suppl 3

7. Sarche M, Spicer P., 2008. Poverty and Health Disparities for American Indian and Alaska Native Children: Current Knowledge and Future Prospects. Ann N Y Acad Sci 1136: 126

8. Tyungu DL, McCormick D, Lau CL, Chang M, Murphy JR, Hotez PJ, Mejia R, Pollack H., 2020. Toxocara species environmental contamination of public spaces in New York City. PLoS Negl Trop Dis 14: e0008249

9. Mejia R, Seco-Hidalgo V, Garcia-Ramon D, Calderón E, Lopez A, Cooper PJ., 2020. Detection of enteric parasite DNA in household and bed dust samples: Potential for infection transmission. Parasit Vectors 13: 1–5

10. Sarink MJ, Koelewijn R, Stelma F, Kortbeek T, van Lieshout L, Smit PW, Tielens AGM, van Hellemond JJ., 2023. An International External Quality Assessment Scheme to Assess the Diagnostic Performance of Polymerase Chain Reaction Detection of Acanthamoeba Keratitis. Cornea 42: 1027–1033

11. Aykur M, Dagci H., 2021. Evaluation of molecular characterization and phylogeny for quantification of Acanthamoeba and Naegleria fowleri in various water sources, Turkey. PLoS One 16

12. Center for New Media & Promotion (CNMP) UCB. My Tribal Area

13. Anon. U.S. Census Bureau QuickFacts: St. George city, Utah. Available at: https://www.census.gov/quickfacts/stgeorgecityutah. Accessed

14. Areekul P, Putaporntip C, Pattanawong U, Sitthicharoenchai P, Jongwutiwes S., 2010. Trichuris vulpis and T. trichiura infections among schoolchildren of a rural community in northwestern Thailand: The possible role of dogs in disease transmission. Asian Biomedicine 4: 49–60

